# Early Lesions Seen in Colonoscopic Surveillance Biopsies from Regions where Colorectal Carcinoma Developed in Patients with Inflammatory Bowel Diseases are Often Not Overtly Dysplastic

**DOI:** 10.64898/2025.12.07.25341783

**Authors:** Xianyong Gui, Konstantin Koro, Scott D. Lee

## Abstract

**BACKGROUND & AIMS:** Screening for precancerous lesions by colonoscopic surveillance in patients with longstanding inflammatory bowel disease (IBD) has been a standard practice. However, failure of detecting precursors and preventing colorectal carcinoma (CRC) development is still common. This study aims to assess whether some lesions shown in the surveillance biopsies eluded our attention due to atypical histopathology.

**METHODS:** 91 patients (M 67/F 24, UC 59/CD 32) with surgically resected IBD-CRC were retrieved from 2 institutions in North America over 20 years. 40 patients with ≥1 colonoscopies performed >1 year prior to cancer diagnosis were regarded as “surveillanced”. Pathology records of CRC resections and all prior colonoscopic biopsies were reviewed.

**RESULTS:** 87.5% of these patients had ≥1 lesion detected in prior biopsies. *Index lesion*, the first lesion identified in all the biopsies, was detected in 65% of patients and in 86.8% of the regions where carcinoma developed later, in which 38.2% were conventional low-grade dysplasia (LGD), 29.4% indefinite for dysplasia (IND), 29.4% inflammatory polyp (IP), and 2.9% serrated lesions (SL).

**CONCLUSION:** In patients who had surveillance, certain lesions were detected by endoscopic biopsies in >80% of patients. >60% of index lesions were not overtly dysplastic, including IND, IP and SL, which may in part represent colitis-associated non-conventional dysplastic lesions.

## INTRODUCTION

Patients with longstanding inflammatory bowel diseases (IBD) have a higher risk of colorectal carcinoma (CRC), due to extensive genetic alterations induced by chronic active inflammation, which results in development of precancerous/dysplastic epithelial changes (mucosal lesions) [1]. Early detection and removal of the precursor lesions are expected to reduce cancer risk. Colonoscopic surveillance, *i*.*e*., periodic colonoscopy with random and/or targeted biopsies to detect mucosal lesions, has been considered as an effective intervention and become a standard practice worldwide for decades [2,3]. However, even in patients who comply with surveillance the occurrence of colorectal carcinoma is still not eliminated. Some studies even found no significant reduction of CRC in IBD patients over the past 40 years despite of colonoscopic surveillance [4]. These findings raise a question about whether some of the precursor lesions present in surveillance biopsies were actually overlooked, especially in view of the recently recognized non-conventional dysplastic lesions in IBD [5-7]. In the present study, we retrospectively reviewed the longitudinal pathology records of the patients with IBD-associated CRC (IBD-CRC) encountered at two institutions in North America over the past two decades.

## MATERIALS AND METHODS

### Study Subjects

We retrieved all patients with surgically resected IBD-CRC at two university-affiliated tertiary medical centers with specialized IBD clinics in the United States and Canada, including the University of Washington Medical Center (UWMC) (Seattle, WA) and the University of Calgary Foothills Medical Centre (UCFMC) (Calgary, AB), over 20-year period (1997 to 2017 for UCFMC, and 1999 to 2019 for UWMC). All the UCFMC patients and the majority of UWMC patients had received care at the same hospital throughout their IBD disease courses.

This study was approved by the University of Washington Institutional Review Board (STUDY00008761) and the University of Calgary Human Research Ethics (HREBA.CC-17-0127).

### Endoscopic and Pathological Studies

The entire histopathology records of the CRC resections as well as prior colonoscopic biopsies, if there were any, were reviewed. For a minority of UWMC patients who had been seen elsewhere prior to surgery, their prior biopsy slides were re-reviewed upon the patients’ transfer, and the pathology diagnoses were documented and available for this study. Otherwise, no additional re-review of histological slides was performed, except for a small subset of biopsies due to ambiguous description in the original reports. The study was approved by the institutional review board at both institutions.

In this study, for those who had at least one colonoscopy with segmental random biopsies or/and targeted biopsies of visible mucosal lesions performed at least one year prior to CRC diagnosis were arbitrarily regarded as “surveillanced” (SURV) ones, while those who never had colonoscopic biopsies since diagnosis of IBD till diagnosis of CRC were regarded as “non-surveillanced” (NSURV) subgroup. For each patient, the pathological characteristics of colorectal carcinomas and findings in all prior colorectal biopsies were extracted from the original pathology reports, with a focus on the mucosal changes/lesions found in the bowel region(s)/segment(s) compared to the cancer site(s). The reported mucosal lesions, *i*.*e*., histologic changes other than inflammation and frank reactive changes, identified on the endoscopic biopsies were reclassified into low-grade dysplasia (LGD, conventional/adenomatous epithelial dysplasia, including diagnosis of “adenoma”), indefinite for dysplasia (IND), inflammatory (pseudo)polyp (IP), and serrated lesion (SL, which was roughly defined in this study and included original diagnoses of sessile serrated adenoma/polyp, serrated lesion unclassified, hyperplastic polyp, and hyperplastic change). The biopsy sites were classified into common anatomic segments of colorectum (*i*.*e*., cecum, ascending, hepatic flexure, transverse, splenic flexure, descending, sigmoid, and rectum). For those originally labeled by the distance (cm) from anal verge, the biopsy locations were reassigned to corresponding bowel segments in conjunction with the endoscopy descriptions.

For each patient, the first ever lesion(s) identified in a prior biopsy taken from the bowel segment/region same as where the carcinoma developed later was arbitrarily defined as “*index lesion(s)*”. The very last noncancerous lesion (excluding high-grade dysplasia) found in a biopsy taken from the same bowel segment/region immediately prior to carcinoma diagnosis was defined as “*latest lesion*”. The lesions found in biopsies from adjacent area within 10cm from the subsequent carcinoma site or in a bowel segment immediately next to the segment where carcinoma arose later were classified as “*adjacent lesions*”, and the lesions located beyond 10 cm or beyond the immediate next bowel segment were classified as “*distant lesions*”.

For each of the index lesions, the corresponding endoscopic appearance of mucosa at the biopsy site was reviewed based on the original endoscopic description and images, and reclassified into “visible” (*i*.*e*., polypoid or abnormal-looking mucosa) and “invisible” (or flat, *i*.*e*., random biopsy with no lesion identified or only inflammation), according to the latest ECCO guideline.^8^ Only the usual high-definition white-light endoscopy was analyzed, partly because many of the endoscopies were performed prior to introduction of chromoendoscopy.

### Statistical Analysis

Categorical variables were summarized using counts and percentages. Student t test and Chi-squared test were used to analyze the differences between groups. *p* value less than 0.05 was considered as statistically significant.

## RESULTS

In total, we retrieved 91 patients, including 38 (UC 27 / CD 11, including 4 with PSC; with 13 surveillanced) from UWMC and 53 (UC 32 / CD 21, including 3 with PSC; with 27 surveillanced) from UCFMC. Data from the two institutions were combined for further analysis, and the patients’ demographic data and the characteristics of carcinomas are also shown in Table 1.

**Table 1.**
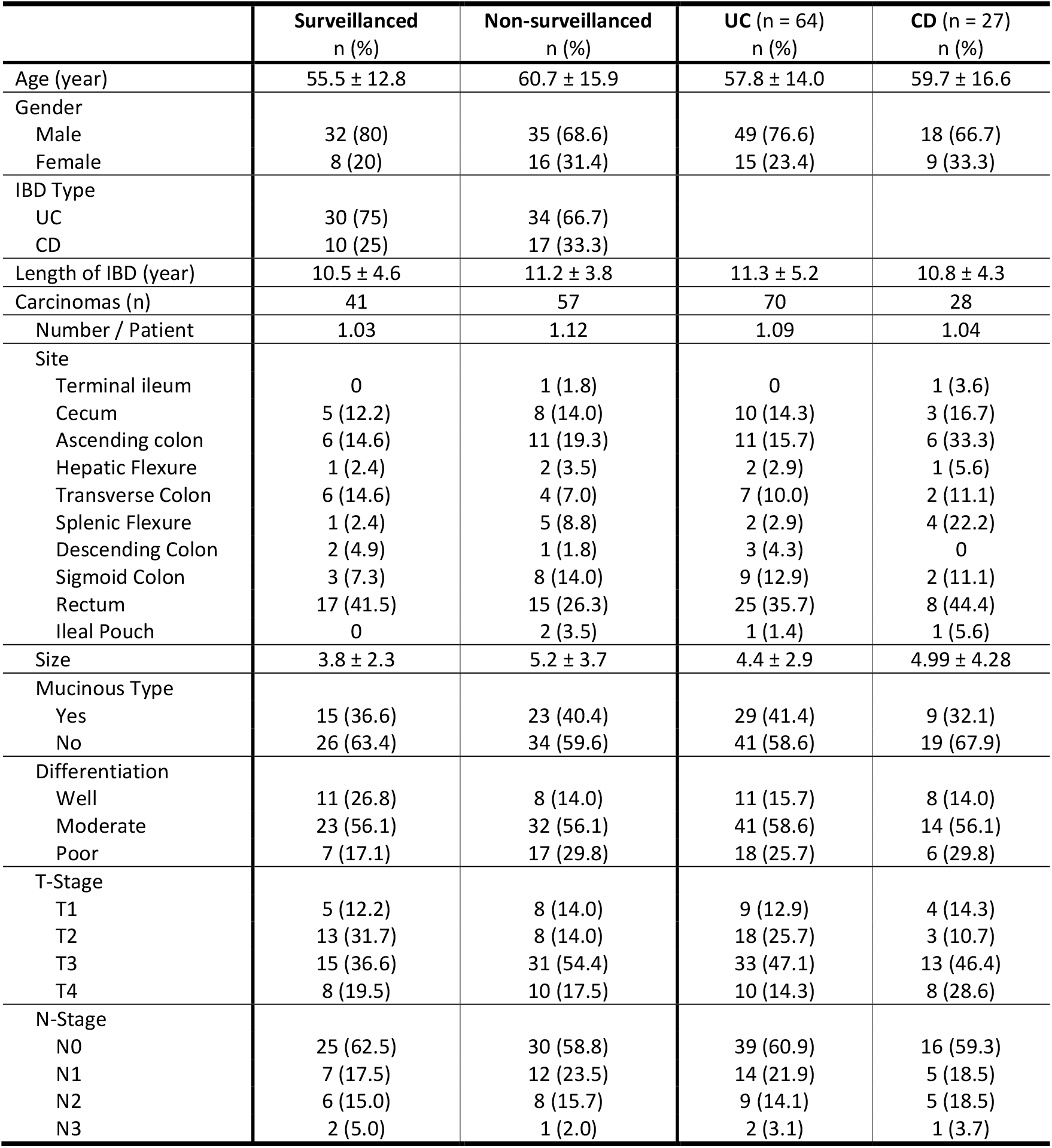
Demographic data of IBD-CRC patients.

|  | Surveilled<br>n (%) | Non-surveilled<br>n (%) | UC (n = 64)<br>n (%) | CD (n = 27)<br>n (%) |
| --- | --- | --- | --- | --- |
| Age (year) | 55.5 ± 12.8 | 60.7 ± 15.9 | 57.8 ± 14.0 | 59.7 ± 16.6 |
| Gender |  |  |  |  |
| Male | 32 (80) | 35 (68.6) | 49 (76.6) | 18 (66.7) |
| Female | 8 (20) | 16 (31.4) | 15 (23.4) | 9 (33.3) |
| IBD Type |  |  |  |  |
| UC | 30 (75) | 34 (66.7) |  |  |
| CD | 10 (25) | 17 (33.3) |  |  |
| Length of IBD (year) | 10.5 ± 4.6 | 11.2 ± 3.8 | 11.3 ± 5.2 | 10.8 ± 4.3 |
| Carcinomas (n) | 41 | 57 | 70 | 28 |
| Number / Patient | 1.03 | 1.12 | 1.09 | 1.04 |
| Site |  |  |  |  |
| Terminal ileum | 0 | 1 (1.8) | 0 | 1 (3.6) |
| Cecum | 5 (12.2) | 8 (14.0) | 10 (14.3) | 3 (16.7) |
| Ascending colon | 6 (14.6) | 11 (19.3) | 11 (15.7) | 6 (33.3) |
| Hepatic Flexure | 1 (2.4) | 2 (3.5) | 2 (2.9) | 1 (5.6) |
| Transverse Colon | 6 (14.6) | 4 (7.0) | 7 (10.0) | 2 (11.1) |
| Splenic Flexure | 1 (2.4) | 5 (8.8) | 2 (2.9) | 4 (22.2) |
| Descending Colon | 2 (4.9) | 1 (1.8) | 3 (4.3) | 0 |
| Sigmoid Colon | 3 (7.3) | 8 (14.0) | 9 (12.9) | 2 (11.1) |
| Rectum | 17 (41.5) | 15 (26.3) | 25 (35.7) | 8 (44.4) |
| Ileal Pouch | 0 | 2 (3.5) | 1 (1.4) | 1 (5.6) |
| Size | 3.8 ± 2.3 | 5.2 ± 3.7 | 4.4 ± 2.9 | 4.99 ± 4.28 |
| Mucinous Type |  |  |  |  |
| Yes | 15 (36.6) | 23 (40.4) | 29 (41.4) | 9 (32.1) |
| No | 26 (63.4) | 34 (59.6) | 41 (58.6) | 19 (67.9) |
| Differentiation |  |  |  |  |
| Well | 11 (26.8) | 8 (14.0) | 11 (15.7) | 8 (14.0) |
| Moderate | 23 (56.1) | 32 (56.1) | 41 (58.6) | 14 (56.1) |
| Poor | 7 (17.1) | 17 (29.8) | 18 (25.7) | 6 (29.8) |
| T-Stage |  |  |  |  |
| T1 | 5 (12.2) | 8 (14.0) | 9 (12.9) | 4 (14.3) |
| T2 | 13 (31.7) | 8 (14.0) | 18 (25.7) | 3 (10.7) |
| T3 | 15 (36.6) | 31 (54.4) | 33 (47.1) | 13 (46.4) |
| T4 | 8 (19.5) | 10 (17.5) | 10 (14.3) | 8 (28.6) |
| N-Stage |  |  |  |  |
| N0 | 25 (62.5) | 30 (58.8) | 39 (60.9) | 16 (59.3) |
| N1 | 7 (17.5) | 12 (23.5) | 14 (21.9) | 5 (18.5) |
| N2 | 6 (15.0) | 8 (15.7) | 9 (14.1) | 5 (18.5) |
| N3 | 2 (5.0) | 1 (2.0) | 2 (3.1) | 1 (3.7) |

In all patients, the bowel regions with carcinoma were involved by colitis in the background (81.5% active, 18.5% quiescent). The average number of colonoscopy-biopsy procedures in SURV group was 5.3 per patient (5.3 ± 3.3, 1 to 15) and 0.5 per year per patient (*i*.*e*., approximately once every 2 years). The average number of biopsies taken for each patient was 11.4 (11.4 ± 4.95, 1 to 16) per colonoscopy and 35 (35.03 ± 32.9, 1 to 163) in total over the years prior to cancer diagnosis. 87.5% (35/40) patients had at least 1 (1 to 15) positive finding (*i*.*e*., detection of some type of mucosal lesions / epithelial changes) in 48.2% (105/218) of the total colonoscopy-biopsy procedures. In 26 patients (74.3% of the surveillance- positive patients and 65.0% of the entire SURV group), 33 index lesions were identified at an interval of 49.7 ± 48.4 (1 to 192) months. The detection rate of index lesions was 86.8% (33/38) of all the corresponding carcinoma sites, and 1.1 lesions per carcinoma (33/29). In 6 cases, the index lesion for each carcinoma site contained mixed two different types of histologic features in either the same mucosal lesion or in different tissue fragments (including fragmented pieces) of a biopsy.

Of all the index lesions, 38.2% were LGD, 29.4% IND, 29.4% IP, and 2.9% SL (Figure 1-A). 7 of the index lesions were available for histopathological review, in which 4 were originally diagnosed as IND and 3 as IP. Upon review, 3 IND and 2 IP were found to have features of crypt dysplasia (*a*.*k*.*a*., dysplasia with terminal differentiation), 1 IND and 1 IP showed marked epithelial serration suggestive of serrated epithelial changes (SEC), 1 IND and 1 IP showed extensive hypermucinous changes, and 1 IP had a small focus of low-grade dysplasia. Representative histomorphologic features are demonstrated in Figure 2.

**Figure 1.**
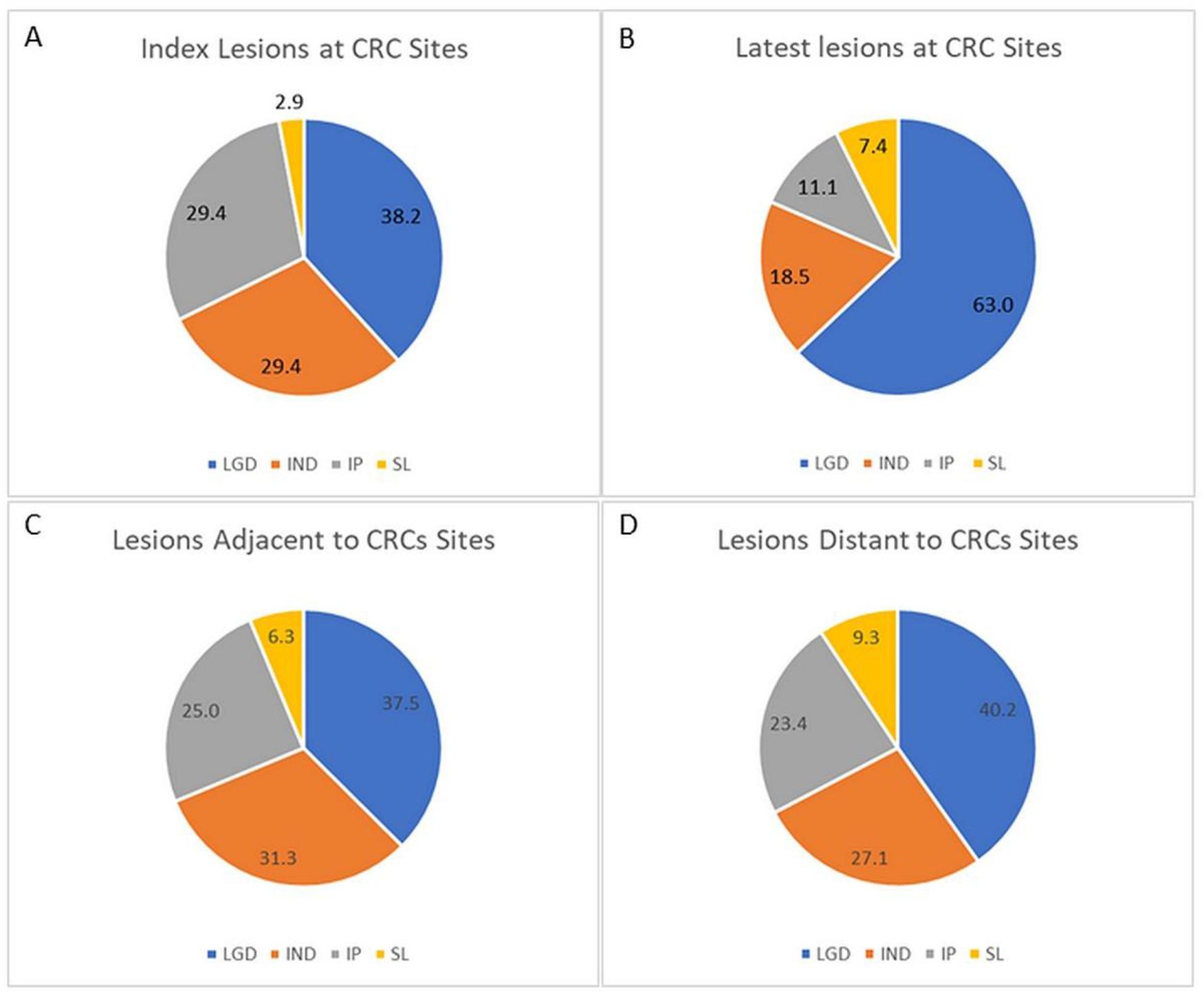
Mucosal lesions identified in previous endoscopic surveillance biopsies. A. Index lesions. B. Latest lesions. C. Lesions adjacent to (<10cm away from) colorectal carcinoma. D. Lesions distant to (>10cm away from) colorectal carcinoma.

**Figure 2.**
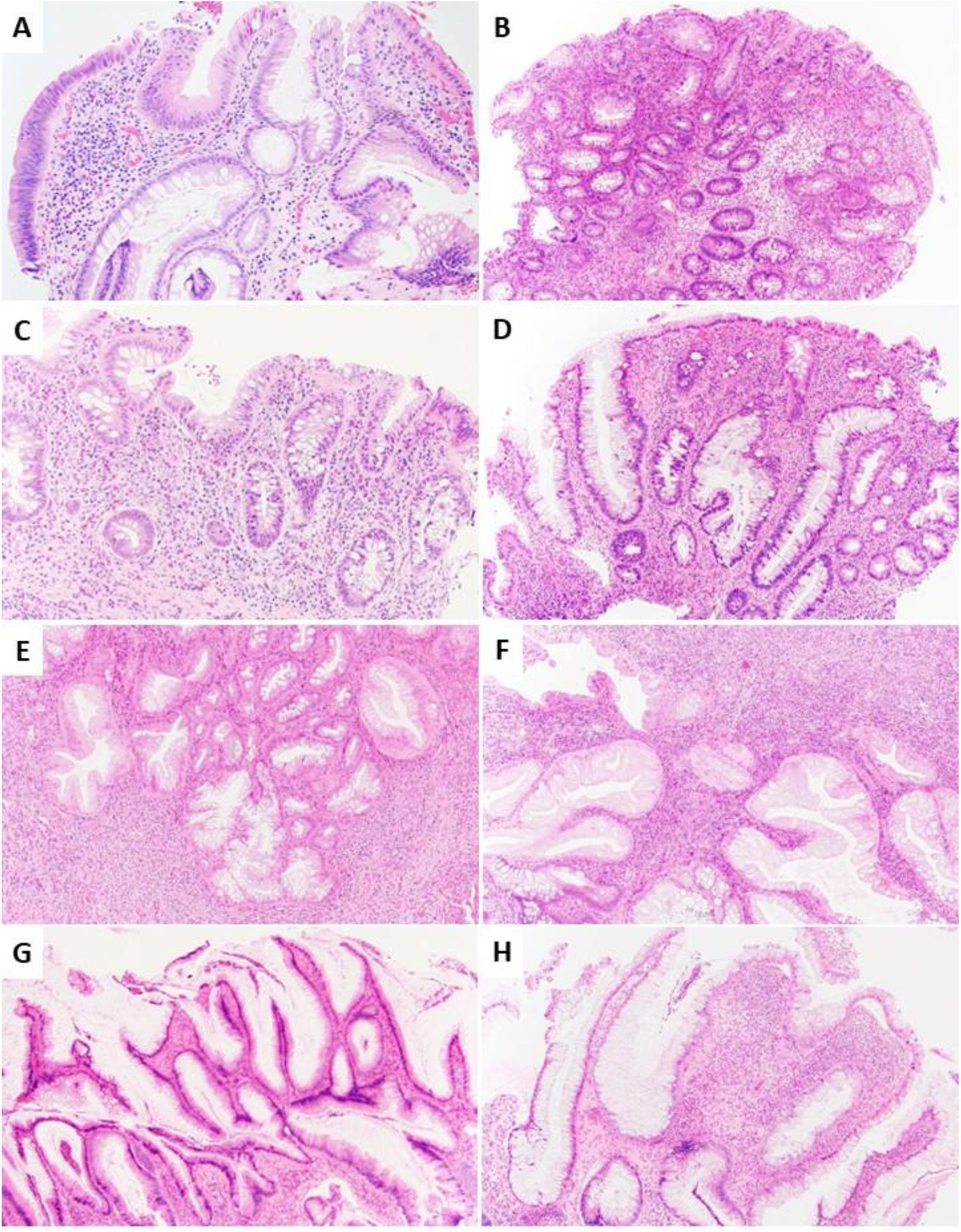
Histomorphologic features seen in some inflammatory polyps. A. A lesion interpreted as indefinite for dysplasia (IND) in which focal epithelial atypia is suggestive but not definitively diagnostic of low-grade dysplasia. B. A small focus of low-grade dysplasia involving both deep crypts and surface epithelium found in an otherwise inflammatory polyp (IP). C/D. Marked cytologic atypia of the deep crypts, compatible with crypt dysplasia, seen in a lesion interpreted as IND (C) and one as IP (D). E/F. Serrated epithelial changes in two IPs. G/H. Hypermucinous epithelial changes seen in an IND (G) and in IP (H).

Following the index lesions, additional lesions were detected later more than once from the same region, in which more than half were LGD (56.1%), followed by IP (29.3%), IND (7.3%) and SL (7.3%).

Upon review of the original endoscopic description about the index lesion biopsy sites, 55.9% (19/34) showed no visible mucosal lesions on white-light colonoscopy images, and 44.1% showed visible lesions with mostly being polypoid and some being non-polypoid irregularities. As outlined in Table 2-A, only a third of the visible lesions were LGD, while the majority showed no overt dysplasia, including IP (46.7%) and IND (20%). Of the invisible lesions, 42.1% were LGD, followed by IND (36.8%), IP (15.8%) and SL (5.3%). On the other hand, 61.5% of LGD and 70% of IND were invisible. Interestingly, of all the lesions, there were equal number of visible and invisible lesions, in which LGD accounted for 42.2%, IP 25.7%, IND 24.8%, and SL 7.4%.

**Table 2.** Mucosal Lesions Identified in Surveillance Colonoscopic Biopsies.

|  | **LGD**<br>n (%)<sup>a</sup> (%)<sup>b</sup> | **IND**<br>n (%)<sup>a</sup> (%)<sup>b</sup> | **IP**<br>n (%)<sup>a</sup> (%)<sup>b</sup> | **SL**<br>n (%)<sup>a</sup> (%)<sup>b</sup> |
| --- | --- | --- | --- | --- |
| **A. Index Lesions** |  |  |  |  |
| Visible | 5 (33.3) (38.5) | 3 (20.0) (30.0) | 7 (46.7) (70.0) | 0 |
| Invisible | 8 (42.1) (61.5) | 7 (36.8) (70.0) | 3 (15.8) (30.0) | 1 (5.3) (100) |
| **B. Latest Lesions** |  |  |  |  |
| Visible | 10 (71.4) (58.8) | 1 (7.1) (20.0) | 1 (7.1) (33.3) | 2 (14.3) (100) |
| Invisible | 7 (53.8) (41.2) | 4 (30.8) (80.0) | 2 (15.4) (66.7) | 0 |
| **C. Adjacent Lesions** |  |  |  |  |
| Visible | 9 (36.0) (50.0) | 2 (8.0) (13.3) | 12 (48.0) (100) | 2 (8.0) (66.7) |
| Invisible | 9 (39.1) (50.0) | 13 (56.5) (86.7) | 0 | 1 (4.3) (33.3) |
| **D. Distant Lesions** |  |  |  |  |
| Visible | 19 (35.8) (44.2) | 9 (17.0) (31.0) | 22 (41.5) (88.0) | 3 (5.7) (30.0) |
| Invisible | 24 (44.4) (55.8) | 20 (37.0) (69.0) | 3 (5.6) (12.0) | 7 (13.0) (70.0) |
| **E. Post-Index Lesions** |  |  |  |  |
| Visible | 9 (40.9) (39.1) | 0 | 10 (45.5) (83.3) | 3 (13.6) (100) |
| Invisible | 14 (73.7) (60.9) | 3 (15.8) (100) | 2 (10.5) (16.7) | 0 |
| **F. Total Lesions** |  |  |  |  |
| Visible | 42 (36.5) (43.3) | 14 (12.2) (24.6) | 51 (44.3) (86.4) | 8 (7.0) (47.1) |
| Invisible | 55 (47.8) (56.7) | 43 (37.4) (75.4) | 8 (7.0) (13.6) | 9 (7.8) (52.9) |

(%)<sup>a</sup>: % of all types of lesions (in visible or invisible category); (%)<sup>b</sup>: % of visible and invisible in the same type of lesions

Of all the latest lesions, 63.0% were LGD, 18.5% IND, 11.1% IP, and 7.4% SL. There was significantly more LGD in the latest lesions as compared to the index lesions (Figure 1-B). Additionally, various types of lesions were also detected in the adjacent and distant regions of bowel, as shown in Table 2-B/C and Figure 1-C&D. Finally, 27 noncancerous lesions were found in the latest biopsies from the same regions (*i*.*e*., latest lesions) at 36.1 ± 44.4 (1 to 192) months prior to carcinoma diagnosis.

## DISCUSSION

The most notable finding of the present study is the identification of earlier mucosal lesions with no overt dysplasia as the presumed index lesions found in a significant population of patients who developed carcinoma later in the same bowel region. Although the lesions may not really be the precursors of the carcinoma, they most likely represent what has happened there. Given the traditional concept that IBD-CRC arises from IBD-associated dysplasia, the surveillance aims at identifying dysplastic lesions, *i*.*e*., unequivocally neoplastic epithelial changes that are similar to adenomatous changes.

Our study identified that about 30% of the index lesions were diagnosed as IND. This observation is in line with a report from Mount Sinai Hospital that IBD patients with IND in surveillance biopsies had a significantly higher (6.85-fold) risk of carcinoma or high-grade dysplasia and was therefore an independent adverse predictor [8]. Moreover, we found that IND was mostly classified as invisible (*i*.*e*., less often polypoid), which is also in agreement with the Mount Sinai findings. Upon re-review in selected cases, some of the IND lesions were found to have histologic features similar to the recently outlined non-conventional dysplasia [5,6].

Another interesting finding of our study is regarding the significance of inflammatory polyp (*a*.*k*.*a*., inflammatory pseudopolyp or postinflammatory polyp), which was also seen in about 30% of the index lesions. IPs are common in both UC and CD, seen in about 20-40% of the patients [9]. Whether IPs have potential of neoplastic transformation has been a lingering question. It has been suggested by some studies that IPs *per se* do not develop into carcinoma but may serve as a surrogate for areas with prior severe inflammation and thus IPs do not independently predict subsequent colorectal neoplasia [9-11]. Some other studies, however, demonstrated an approximately 2-fold increased risk of CRC in patients with IPs and therefore suggested IP as a risk factor of colorectal neoplasia [12-15], which lead guidelines to recommend intensifying surveillance to patients with multiple IPs [2]. Histologically, there are several forms of IPs. Some are mucosal protuberance composed largely of granulation tissue as a result of remained necroinflamed mucosal island(s) in between deep ulcers, and some are redundant mucosal tags or projections composed of intact regenerative mucosa with or without active inflammation [16-18]. Some IPs may contain a mild reactive cytologic atypia or even focally mixed dysplasia, and they often display concurrent serrated/hyperplastic features or hypermucinous or eosinophilic (goblet cell-depleted) epithelial changes, which are all the features of the recently recognized non-conventional dysplasia in IBD [19]. In addition, many of the true colitis-associated dysplastic lesions show morphologic features of IP, often as granulation tissue, in the background [19]. In some cases, IP may mask a less visible adjacent dysplastic lesion. De Cristofaro et al reviewed 105 pseudopolyp-like lesions removed endoscopically in patients with longstanding IBD (80 UC/25 CD). They found that nearly a quarter of the lesions bear dysplastic foci and half show mixed hyperplastic features, more common in large lesions (>5 mm) and those in right colon [20]. With regard to our findings, it is difficult to determine whether IPs were simply as confounders or truly preneoplastic lesions that evolved into neoplastic precursor lesions of subsequent carcinomas. However, based on our data, recommendation of a more aggressive surveillance strategy in patients with the presence of IPs seems to be reasonably justified. In-depth study on potential neoplastic features at molecular level in IPs is also suggested.

We also found that approximately 3% of the index lesions were characterized by serrated or hyperplastic epithelial changes with sawtooth-like surface and/or glandular epithelium. A subset of these serrated lesions fit the diagnostic criteria of sessile serrated adenoma/lesion (SSA/L) characterized by basal crypt dilatation. Some of the lesions appeared to be not significantly different from usual hyperplastic polyps. Some seem to be compatible with the so-called serrated epithelial change (SEC) that has been found to be associated with coexistent or subsequent dysplasia and increased risk of high-grade dysplasia and carcinoma, especially when it is seen on successive endoscopic biopsies [21]. Therefore, SEC has been proposed to be a variant of non-conventional dysplastic lesions in IBD and has been reclassified as serrated dysplasia NOS [5,7].

In line with the well-recognized experience, classic low-grade dysplasia (LGD) was still the most common precancerous lesions in our data, accounting for nearly 40% of the index lesions. The reported rate of progression of LGD to carcinoma varies between studies, with a high progressing rate reported in earlier literature [22] but lower rate in recent reports. In a large nationwide cohort with long-term follow-up of IBD patients with LGD, the cumulative incidence of colorectal advanced neoplasia (CRC or HGD) was 21.7% after 15 years [23]. Lighter et all reported that 7% of patients with detected LGD eventually developed advanced neoplasia after a median period of 86 months, and the overall cumulative incidence of CRC at 10 years after initial diagnosis of LGD was 8.5% [24]. Another recent study from Cleveland Clinic found that the 1- and 2-year cumulative rate of progression from LGD to carcinoma were 0.7% and 1.6%, respectively [25]. Older age of patients, large (>1cm) or flat LGD were found to be more likely to progress [26,27]. Between UC and CD, the rate of progression from LGD to CRC was similar [28].

In the literature, there is inconsistent reporting about whether the dysplastic lesions are more often visible or invisible [29,30]. Invisible LGD was found to have a 2-fold higher risk of progression than visible ones [25,26]. In our data (Table 2), the LGD seen in the index lesions were mostly (>60%) invisible by white-light endoscopy. Interestingly, when taken together, the overall LGD lesions seen at different times and in different bowel regions in our patients, the chance of a dysplastic lesion being visible or invisible was almost equivalent. Certainly, the variation in rates of LGD visibility may also be affected by the improvement in endoscopic visualization over time, not to mention the current wide application of chromoendoscopy.

Location discordance of carcinoma and precancerous lesions has been a common observation, that in part may reflect multifocality of mucosa at risk of neoplastic transformation in patients with IBD. Although our study limited the index lesion as those located in a bowel segment concordant to subsequent carcinoma, it is not possible to determine whether the carcinoma site was exactly the original biopsy spot of the index lesion. It is also impossible for us to assess whether the index lesions were removed completely and whether any possible remainder of a lesion was truly the precursor of the subsequent carcinoma in any given patient. In addition, we noticed many similar lesions were also present simultaneously in some other regions of bowel, both adjacent and distant to carcinomas. Whether multifocality of dysplasia increases the risk of carcinoma development as compared to unifocal lesion was inconclusive, as studies so far reported opposing results [25,26,31].

In our data, the compositions of previous lesions in UC and CD did not show significant difference, which is in line with the consensus that the risk of CRC in UC and in CD is similar [28].

Two patients in this series, including one initially diagnosed as UC but later changed to CD, developed carcinoma in ileal pouch. Both were not under endoscopic surveillance and had no known clinical history of primary sclerosing cholangitis (PSC). Given this finding, patients treated with colectomy and ileal pouch-anal anastomosis (IPAA) should also continue surveillance, especially for those with PSC and history of refractory pouchitis, as suggested by ASGE [32]. A Netherlands study on IBD patients with IPAA over 20 years found 1.8% of the patients developed dysplasia and 1.3% developed carcinoma in pouch [33].

In summary, our study demonstrates that in IBD patients who developed enteric carcinoma, the precancerous/preneoplastic lesions arose in the same areas were detected in more than 80% of patients prior to cancer diagnosis, and more than half of the lesions were not overtly dysplastic. We speculate that some of the lesions may represent a potential inciting precursors from which the carcinoma arose, and at least some of these lesions may in fact be the colitis-associated nonconventional dysplastic lesions. Whether these early lesions were indeed the ancestor lesions that progressed into carcinomas, or they were not direct precursors but simply reflected the underlying cancer-driven genetic alterations is a question that we cannot answer. Future genomic study, including phylogenetic assessment, to clarify the possible association of these index lesions with the colorectal carcinomas is awaited.

Admittedly, there are several limitations in this study. First, the retrospective nature of our data based on the reported original pathology and endoscopic descriptions is acknowledged. Not all archival pathology materials undergone second review for this study. Second, some of the diagnoses were reported in the days when the non-conventional dysplasia in IBD was not yet known. Third, the completeness of endoscopic removal of the earlier lesions could not be assessed with certainty. Fourth, the determination of visible and invisible lesions was made on white light endoscopy prior to chromoendoscopy was available, and thus may not be representative by the latest standard of chromoendoscopy and other high-definition endoscopic technologies.

## Data Availability

The data underlying this article cannot be shared publicly for the privacy of individuals that participated in the study. The data will be shared on reasonable request to the corresponding author.

## Author Approval

All authors have seen and approved the manuscript.

